# Variation in human mobility and its impact on the risk of future COVID-19 outbreaks in Taiwan

**DOI:** 10.1101/2020.04.07.20053439

**Authors:** Meng-Chun Chang, Rebecca Kahn, Yu-An Li, Cheng-Sheng Lee, Caroline O. Buckee, Hsiao-Han Chang

## Abstract

**Background:** As COVID-19 continues to spread around the world, understanding how patterns of human mobility and connectivity affect outbreak dynamics, especially before outbreaks establish locally, is critical for informing response efforts. In Taiwan, most cases to date were imported or linked to imported cases.

**Methods:** In collaboration with Facebook Data for Good, we characterized changes in movement patterns in Taiwan since February 2020, and built metapopulation models that incorporate human movement data to identify the high risk areas of disease spread and assess the potential effects of local travel restrictions in Taiwan.

**Results:** We found that mobility changed with the number of local cases in Taiwan in the past few months. For each city, we identified the most highly connected areas that may serve as sources of importation during an outbreak. We showed that the risk of an outbreak in Taiwan is enhanced if initial infections occur around holidays. Intracity travel reductions have a higher impact on the risk of an outbreak than intercity travel reductions, while intercity travel reductions can narrow the scope of the outbreak and help target resources. The timing, duration, and level of travel reduction together determine the impact of travel reductions on the number of infections, and multiple combinations of these can result in similar impact.

**Conclusions:** To prepare for the potential spread within Taiwan, we utilized Facebook’s aggregated and anonymized movement and colocation data to identify cities with higher risk of infection and regional importation. We developed an interactive application that allows users to vary inputs and assumptions and shows the spatial spread of the disease and the impact of intercity and intracity travel reduction under different initial conditions. Our results can be used readily if local transmission occurs in Taiwan after relaxation of border control, providing important insights into future disease surveillance and policies for travel restrictions.

## BACKGROUND

The Coronavirus Disease 2019 (COVID-19) was first reported in Wuhan, China in December 2019 and has since caused a global pandemic, with over 15,000,000 confirmed cases and over 600,000 deaths reported by July 23, 2020.^1^ Scientific discoveries have advanced at an unprecedented pace, with numerous clinical trials of drugs and vaccines underway.^2,3^ In the meantime, public health officials must rely on other interventions, such as social distancing and travel restrictions, to slow the spread and reduce the peak of the outbreak, in order to prevent health systems from being overwhelmed.^4,5^

In January 2020, as the epidemic in Wuhan grew, many countries implemented travel bans, and airlines canceled flights to attempt to slow the spread.^6^ A number of studies have estimated the risk of importation globally, with some suggesting up to two-thirds of all imported cases went undetected.^7,8^ For Taiwan, there have been 411 reported cases as of June 30, 2020 ^9^, with 356 imported (87%) and 55 local cases (13%). 52 local cases (94.5 %) were linked to imported or known cases, and 3 local cases (5.5 %) have unknown origin. Since February 7th, Taiwan has implemented entry restrictions on foreign nationals based on their travel histories; 14-day home quarantine started being required for visitors from certain locations from February 10th and became required for all travelers from March 19th.^10^ While COVID-19 transmission in Taiwan is relatively well-controlled, the number of cases globally is still increasing.^1^ When border control is relaxed in Taiwan in the future, importation from other countries has the potential to lead to local outbreaks, especially if other non-pharmaceutical interventions, such as hand washing or mask wearing, are not adopted at the same level as in March and April 2020.

As the number of cases globally due to community transmission grows relative to the number of imported cases, attention has turned to more local measures to decrease spread, such as cancellations of mass gatherings, business closures, and local travel restrictions.^11^ Mobility data can provide critical information for responding to outbreaks and understanding the impact of travel restrictions.^12^ Recent studies have analyzed the effects of human mobility and travel restrictions on disease spread during the COVID-19 pandemic.^6,13–16^ Here, to prepare for COVID-19 and its impact, in collaboration with Facebook Data for Good, we describe the metapopulation models we’ve built that include human movement data to better understand the high risk areas of disease spread and assess the potential impact of local travel restrictions in Taiwan.

## METHODS

### Movement data and geographic unit

We incorporated two different sources of mobility data from Facebook into our models: Facebook colocation data and Facebook movement data. Facebook’s newly developed colocation matrices (*Facebook colocation data*) give the probability that people from two different geographic units will be in the same 600 m × 600 m location for five minutes using data over the course of a week. Facebook’s regular movement data (*Facebook movement data*) aggregates the number of trips Facebook users make between locations every eight hours (%Figure S1).^17^ Mobility data between January 26^th^ and June 30^th^ were used. Facebook movement data were disaggregated by weekdays (Monday to Friday), weekends, and holidays (Lunar New Year, Ching Ming festival, and Dragon Boat Festival). Facebook colocation data included weeks containing holidays and weeks containing only regular (i.e. non-holiday) days.

The geographic unit used in this study was at the centrally-governed level of “city” (here “city” indicates city, county or special municipality in Taiwan). Shape files were downloaded from Government open data platform (https://data.gov.tw/dataset/7442). We excluded three cities outside of the main island of Taiwan from the analysis due to their low connectivity with the main island, leaving 19 cities.

### Metapopulation Models

We developed susceptible-latent-infectious-recovered (SLIR) models of the spread of COVID- 19 throughout Taiwan. We ran the models stochastically to understand the initial stages of disease spread. We ran the model until either (1) it reached *n* cumulative infections or (2) the total number of infections became 0 to estimate the probability of having more than *n* infections (denoted by *P_n,k_*, where *k* represents the number of initial infections), the time it takes to reach *n* infections (denoted by *T_n,k_*), and the standard deviation of infection numbers at *T_n,k_* (denoted by *V_n,k_*). To assess the initial stages of the outbreak, we used *n*=1000 and *k*=3 as our baseline values.

Let *S_i_, L_i_, I_i_, R_i_* be the number of susceptible, latent, infectious, and recovered individuals in location *i*, respectively, and *N_i_* be the total population in location *i*. Let *D_L_* (=3.5) be the latent period, and *D_I_* (=3) be the duration of infectiousness.^13^ Because transmission rates can change with non-pharmaceutical interventions, as shown in previous studies, we vary *R_0_* in our model (*R_0_* =2.4, 1.2, or 0.9).^13,18,19^

We modified spatial models from previous studies^20-23^ and constructed two metapopulation models, a contact model and a residence model, with the former using Facebook colocation data and the latter using Facebook movement data. In the “contact model”, we assumed that contact rates (and therefore transmission rates) varied among locations and was proportional to colocation probabilities (*C_ij_*, the probability that a person from location *i* collocates with a person from location *j*) from Facebook colocation data. We scaled *R_0_* by *C_ij_*N_j_* (for *j* not equal to *i*) or *C_ii_*\*(*N_i_*−1), standardized to *C_ii_*\*(*N*−1) in Taipei.

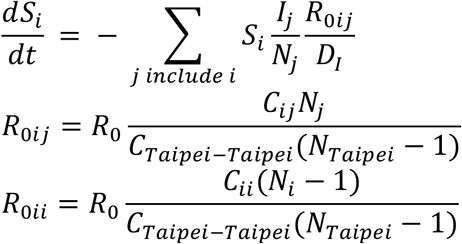

In the “residence model”, we first estimated the proportion of time people living in location *i* spend in location (*P_ij_*) based on Facebook movement data (see details in Supplementary Methods), and modeled the transmission dynamics by considering both that (1) non-travelers get infected by infectious visitors to their home location (the first part in the following equation) and that (2) naïve travelers get infected when they travel (the second part in the following equation).

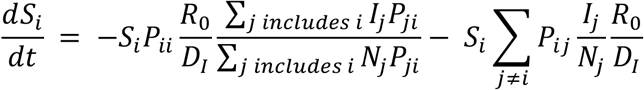

The remaining equations are the same across the two models.

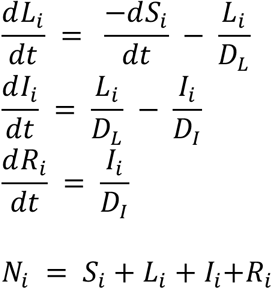

In addition to using different movement data, the major difference between two models is that the transmission rate within each city (*R_0_/D_I_*) varies with colocation matrices in the contact model, while it remains constant in the residence model. In this sense, the contact model is similar to the traditional density dependent model, where contact rates (and therefore transmission rates) vary with population density, and the residence model is similar to the frequency dependent model.^24^ As it is unclear which is most appropriate for COVID-19, we used both and compared the results.

### Risk of infection and regional importation

We defined three connectivity measures relevant for disease transmission, *risk of infection, risk of regional importation*, and *source of importation*. Using Facebook colocation data, we defined *R_0ii_* as intracity *R_0_* and 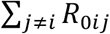 as intercity *R_0_* for location *i*. The sum of intracity *R_0_* and intercity *R_0_* reflects total risk of infection and was standardized to the highest value.

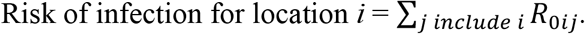

Similarly, using Facebook movement data, we defined 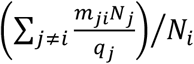 as risk of regional importation (i.e. importation from other cities within Taiwan) for location *i*, where *q_j_* represents the average number of subscribers in location *j* and *m_ji_* represents the average number of people moving from location *j* to location *i* per unit of time in Facebook movement data. Source of importation was defined as the number of travelers from each location *i* and standardized to the highest value.

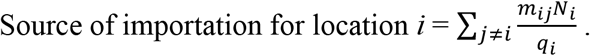

Facebook colocation data from regular days were used to calculate risk of infection, and weekday movement data were used to calculate risk of regional importation and source of importation.

### Modeling travel reduction

To assess the potential effect of travel restrictions at multiple levels, we modeled either intra-city travel reductions, inter-city travel reductions, or a combination of both travel reductions (“overall reduction” in texts and figures) for 1, 2, 3, or 6 months or for the whole period of time. Travel reductions started from the beginning of the simulations or when there were 10, 20, 30, 50, or 100 accumulated infections. The proportion of reduction is denoted by *G*. In the contact model, intracity reduction was modeled by *R_0ii_*\*(1−*G*) for all *i*, and intercity reduction was modeled by *R_0ij_*\*(1−*G*) for all *i* not equal to *j*. In the residence model, intracity reduction was modeled by *R_0_*\*(1−*G*) and intercity reduction was modeled by *P_ij_*\*(1−*G*) for all *i* not equal to *j* and *P_ii_* +(1−*P_ii_*)*\*G* for all *i*.

## RESULTS

### Varying human mobility across space and time in Taiwan

We quantified how intercity and intracity mobility varied at the centrally-governed level in the past five months using Facebook mobility data. On average, intracity movement was 7-fold of intercity movement, and intracity colocation probability was over 200-fold of intercity colocation probability. We quantified the difference in connectivity between locations by three measures: risk of infection, risk of regional importation, and source of importation (Figure 1 and Table S1). Risk of infection identifies locations with larger colocation probabilities. If assuming contact rates were proportional to colocation probabilities, disease transmission rates are expected to be higher in locations with higher risk of infection, such as Taipei City, New Taipei City, and Kaohsiung City. Risk of regional importation represents the relative number of travelers and local people, and higher values indicate higher possibility that travelers will transmit virus to local people. Source of importation calculates how much travelers from each location contribute to other locations. Viruses are expected to spread more quickly if initial local infections in Taiwan occur in locations with higher values of source of infection. Taipei and New Taipei City are cities with the highest risk of regional importation and source of importation, respectively. These three measures quantify different but related aspects of mobility and connectivity that are relevant for disease transmission, and while well-connected cities tend to have high values for all three measures, there are still some differences among them.

**Figure 1.**
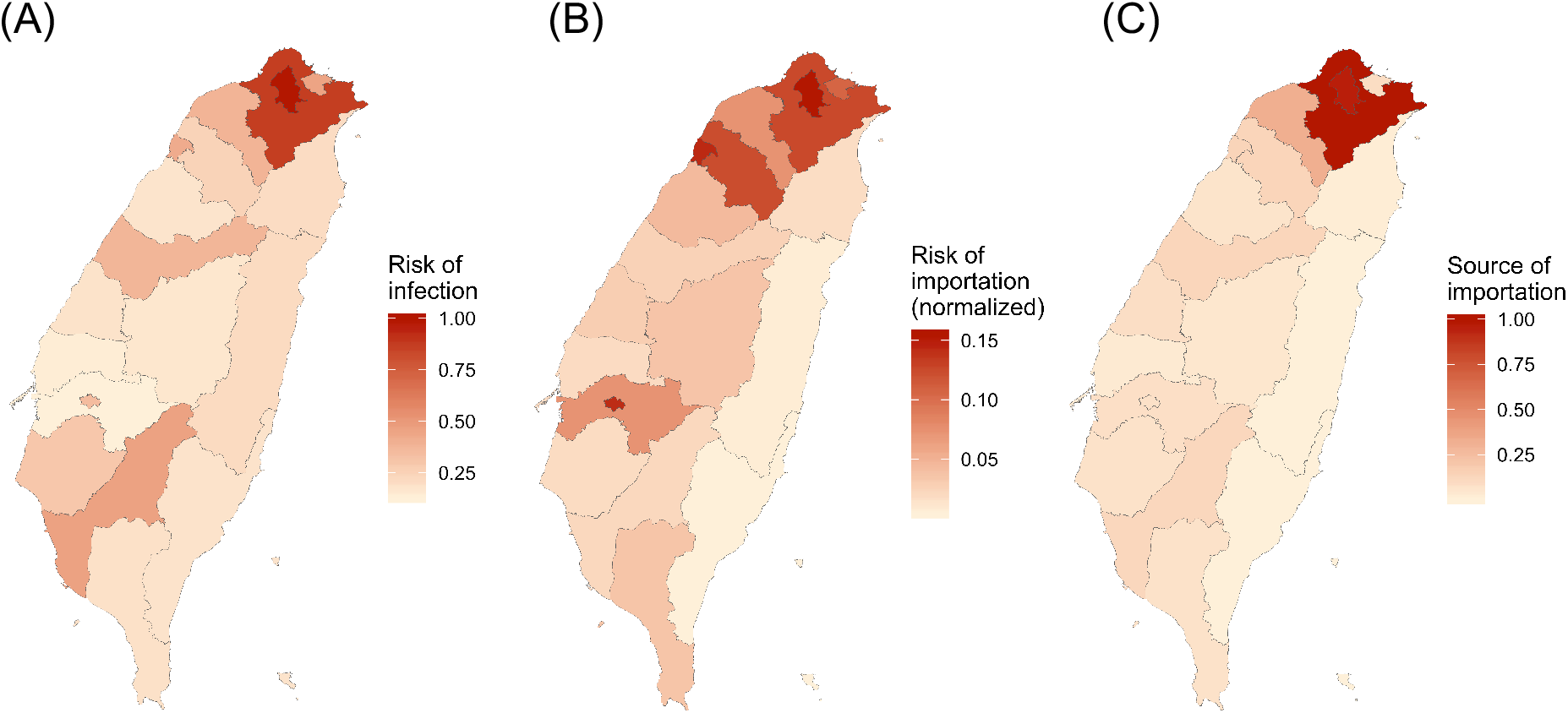
Connectivity measures. Three kinds of connectivity measures relevant to disease spread are shown. The values for bigger cities were larger. (A) Risk of infection. (B) Risk of importation. (C) Source of importation.

We found intercity mobility between some of the locations first decreased and then increased in the past few months, which is consistent with the changes in the number of local cases in Taiwan and global number of cases (%Figure S2 and %Figure S3), indicating the level of change that can happen without travel restrictions imposed by the government. We also observed significant changes during holidays. Lunar New Year was within the early stages of the SARS-CoV-2 outbreak, and for most of cities pairs (95%), colocation probabilities during Lunar New Year was significantly higher than regular days, as expected during holidays. However, the proportion of city pairs with higher than usual intercity connectivity during the Ching Ming Festival, which occurred at the time when the number of cases was increasing dramatically globally and the number of local cases was just starting to decrease, decreased to 67%. Dragon Boat Festival was at the time the number of local cases remained zero for more than a month, and the proportion of city pairs with higher than usual intercity mobility increased to 76%.

### The impact of the location of initial infections on the risk of the spread

At the end of June 2020, most cases in Taiwan were imported or linked to imported cases. Therefore, we used meta-population models parameterized by human mobility data from Facebook to simulate the spread of SARS-CoV-2 under a variety of initial conditions, including both the number of initial infections and their locations. We developed a web-based interface (https://roachchang.shinyapps.io/TW_CoV_Dynamics/) to show the geographic distribution of infections given different initial conditions, which can be readily used to inform targeted surveillance and control if SARS-CoV-2 starts spreading locally in Taiwan. Because other disease-relevant hygiene behaviors, such as hand washing, mask wearing, or social distancing, may also have changed due to the awareness of COVID-19, we explored different transmission rates (*R_0_* =2.4, 1.2, or 0.9).

We considered different aspects of disease spread – the probability of outbreak, the speed of spread, and the geographic range of outbreak. We estimated the probability of having more than 1000 infections (denoted by *P_1000,k_*, where *k* represents the number of initial infections) using stochastic simulations and used this to represent the probability of an outbreak. As expected, we found that, if we assumed that the transmission rates varied among cities (contact model), the probability of having more than 1000 infections also varied with the locations of initial infections, with the cities with larger risk of infection showing larger *P_1000_* (%Figure S4A and Table S2). In simulations where 1000 infections were reached, the time it took to reach 1000 infections (denoted by *T_1000,k_*) was also shorter for cities with larger risk of infection (%Figure S4B). When assuming that the transmission rates in different cities were the same (residence model), the probability of having more than 1000 infections and the time to reach 1000 infections did not vary much with the locations of initial infections (%Figure S5 and Table S3). The effect of intercity connectivity, however, was reflected in the variation in infection numbers across cities at *T_1000_* (denoted by *V_1000_*). The variation in infection numbers was lower in cities with higher values of source of importation (%Figure S4C) as the chance of spreading the virus to other cities was higher. In both models, well connected cities played more important roles, as they spread the virus to other cities more quickly and more widely.

### The impact of varying mobility on the risk of spread

Above results were based on mobility data on regular days. Given that human mobility varied significantly in the past few months without travel restrictions imposed by the government, we further quantified the impact of varying mobility in Taiwan on the risk of spreading SARS-CoV-2. The impact was mainly reflected in the geographic range of infections in both models. When initial infections occurred in or around Lunar New Year, the speed of disease spread was enhanced (Figure 2). Because mobility during Ching Ming Festival and Dragon Boat Festival differed less from regular days and these two holidays only lasted four days, it only led to minor differences in the geographic range of infections (Figure 2). In the contact model, the probability of local outbreak was higher if initial infections occurred in or around Lunar New Year, and this impact was more apparent when initial infections were in locations with lower risk of infection (%Figure S6).

**Figure 2.**
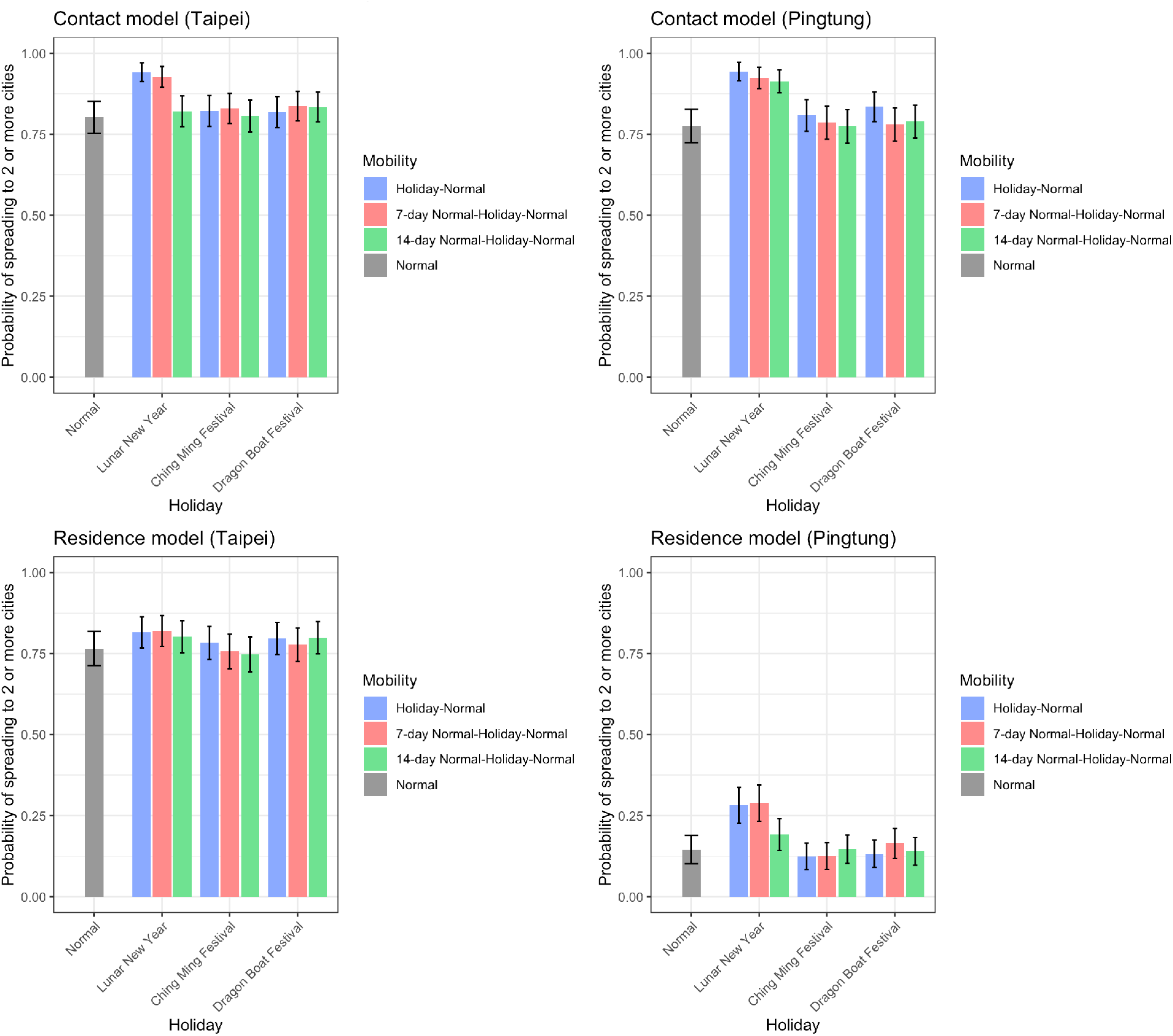
The impact of holiday travel on the disease spread. The speed of disease spread, quantified by the probability of spreading to 2 or more cities when it reaches 50 infections, from simulations with initial infections in Taipei City (representing big cities) or Pingtong County (representing small cities) are shown. The impact of Lunar New Year (10-day) was larger than Ching Ming Festival (4-day) and Dragon Boat Festival (4-day). Initial infections occurred either in (blue) or before holidays (red: 7-day; green: 14-day). *R_0_*=2.4.

### The effect of travel restrictions

We examined the impact of varying mobility that occurred naturally during holidays on the disease spread above. Here we explored the level to which travel restrictions imposed by the government could potentially reduce the spread of SARS-CoV-2 in Taiwan at the initial stage of an outbreak. In both the contact model and the residence model, decreasing intracity movement had a much larger impact on *P_1000,3_* (Figure 3) and *T_1000,3_* (%Figure S7) than decreasing intercity movement. The impact of reducing intercity travel was most evident in influencing how widespread the virus was: the infections were located in only a few cities at *T_1000,3_* if intercity travel was reduced (%Figure S8).

**Figure 3.**
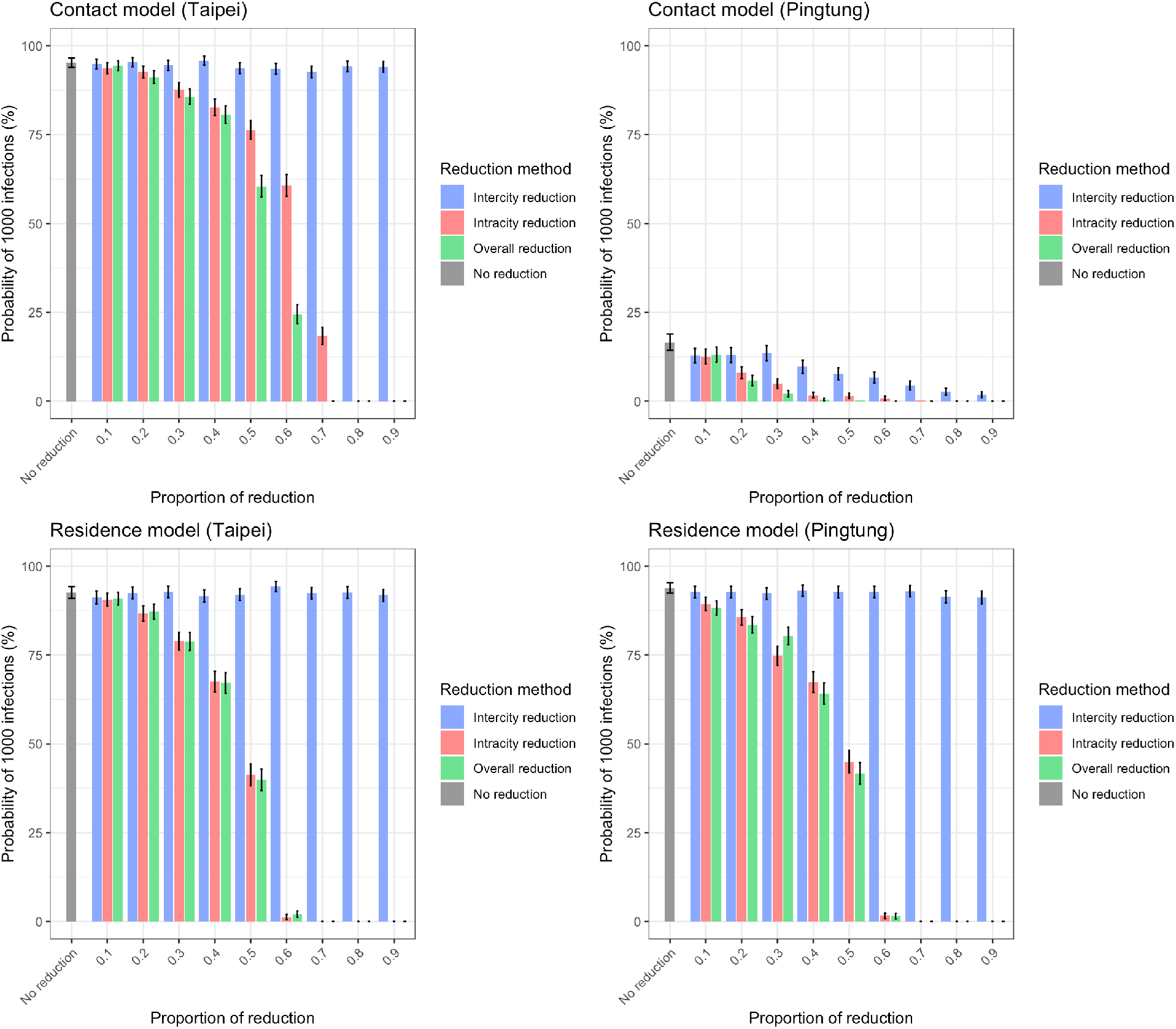
The impact of travel reduction on the probability of having 1000 infections. *P_1000,3_* from simulations with initial infections in Taipei City (representing big cities) or Pingtong County (representing small cities) using both contact and residence models are shown. The difference between big and small cities was more significant in the contact model than in the residence model. Intracity and intercity travel reduction reduced *P_1000,3_*, while the impact of intercity travel reduction was minor. Here travel reduction was applied during the whole time and *R_0_*=2.4.

We then investigated the impact of duration and timing of travel reductions (Figure 4, and details at https://roachchang.shinyapps.io/TW_CoV_Dynamics/). The probability of local outbreak decreased with increased duration of intracity travel reduction, but not change with the duration of intercity travel reduction. The results suggest that higher levels of reduction and longer periods of reduction for intracity travel can have similar impacts. For example, a 60% intracity travel reduction for 20 days had similar outcomes as a 70% reduction for 10 days. While *P_1000,3_* did not change with the length of intercity travel reduction, longer intercity travel reduction led to slower progression of the outbreak (higher *T_1000,3_*) in the contact model and more clustered infections (higher *V_1000,3_*) in both models (%Figure S9). Furthermore, among the parameters we used, it was the best to reduce travel as early as possible to reduce the risk of outbreak (%Figure S10).

**Figure 4.**
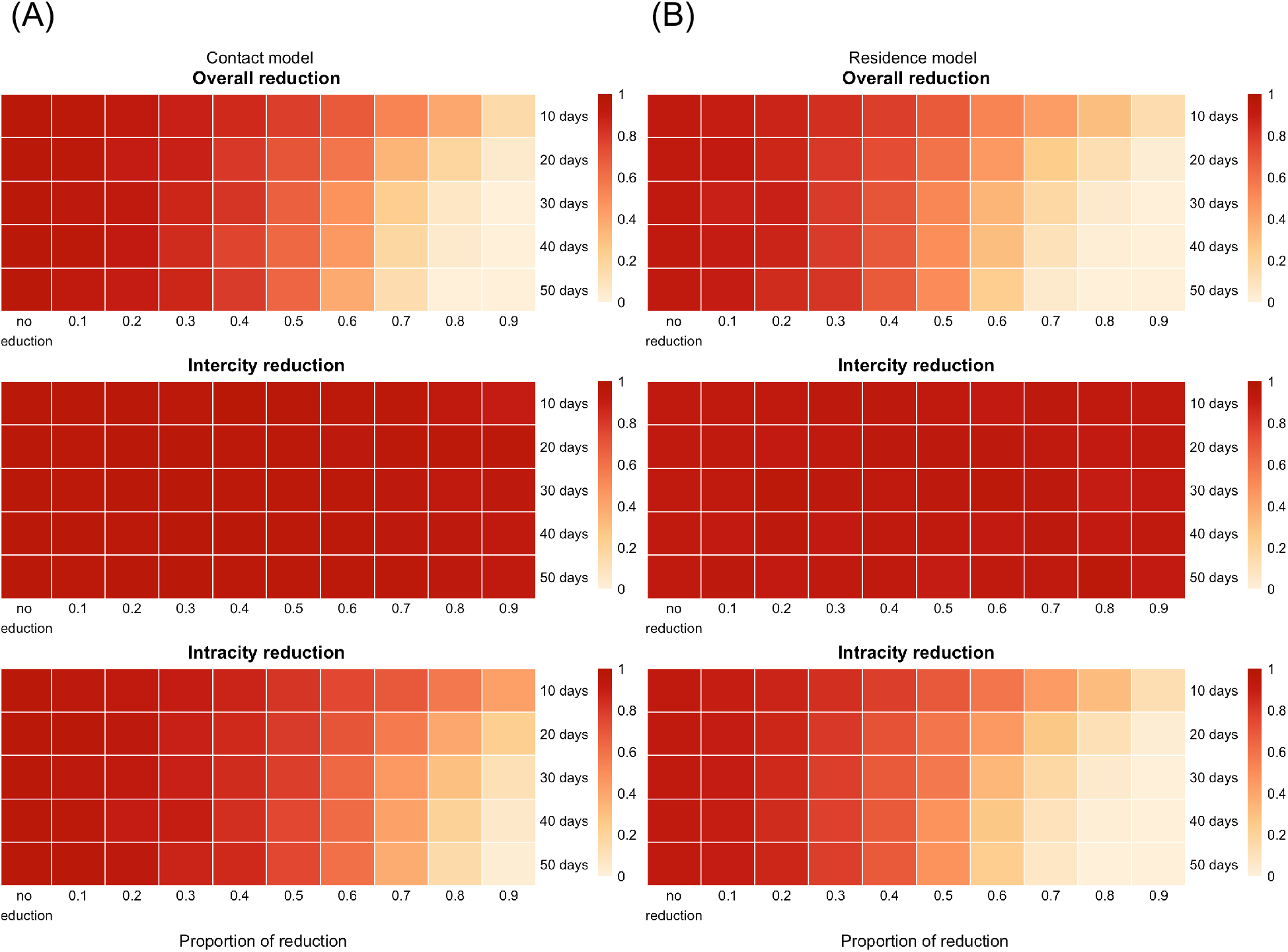
The impact of the duration of travel reduction and the level of reduction on the probability of having 1000 infections. *P_1000,3_* from the contact model (A) and the residence model (B) with initial infections in Taipei City and *R_0_*=2.4. The color represents the level of reduction in *P_1000,3_* (white to red represents smaller to larger reduction). As the duration of intracity travel reduction increased, *P_1000,3_* decreased in both models. *P_1000,3_* did not change with the duration of intercity travel reduction.

## DISCUSSION

By utilizing aggregated human mobility data from Facebook, we characterized how mobility patterns in Taiwan changed since the emergence of COVID-19, and built metapopulation models to understand the potential spread of SARS-CoV-2 in Taiwan and assess the potential impact of travel restrictions. We identified the top cities with the highest risk of infection as well as the top cities with the highest importation risk from other cities based on Facebook data and population sizes. We made a web-based interface showing the geographic distribution of infections at different time points (*T_100_, T_500_* and *T_1000_*) in the initial stages of the outbreak given different locations of initial infections. We demonstrate that these modeling results based on empirical mobility data can be obtained before an outbreak occurs, and can be readily used to help the public avoid high-risk areas, help public health professionals identify surveillance targets, and inform decisions on travel restrictions, providing one of the key elements for COVID-19 preparedness.

Consistent with previous findings showing that international or domestic travel bans are less effective than social distancing,^6,25^ we found that intracity travel reduction has a higher impact on disease dynamics than intercity travel reduction and increasing the length of intracity travel reduction increases the impact. Intercity travel reduction, however, influences the variation in infection numbers across cities and can reduce the number of cities that have infections at the initial stage of the outbreak. While intercity travel did not decrease the probability of outbreak, containing the infections to a few cities has important public health impacts, as this means surveillance system can focus on fewer cities and control efforts can be more targeted. Practically, intercity reduction might be easier to implement than intracity reduction.

Intracity travel reduction in our model is effectively the same as any measure that can reduce contact rates between individuals, such as social distancing, or transmission probability given contact, such as hand washing or wearing facemasks. These measures have been shown to be effective in reducing the transmission of respiratory viral pathogens in both modeling and empirical studies,^26-30^ and should be encouraged. It has been shown that contact rates can be reduced by more than 70% during a lockdown.^31^

Our study found that similar probabilities of outbreak can occur with various combinations of length, level, and timing of travel reductions. Health officials can therefore take into consideration feasibility of different interventions, impact on society, and the capacity of the healthcare system to determine the optimal interventions and their duration.^5^ Because the volume of travel in and around holidays can increase the speed of virus spread, our results suggest that it is important to avoid travel or reduce the impact of travel through measures such as limiting social interactions and wearing facemasks when taking public transportation to reduce the spread of the virus.

We showed that Facebook mobility data can be used to track how the volume and pattern of travel change through time as the outbreak progresses, and we can incorporate any change in human mobility into the metapopulation models in nearly real time to help fight COVID-19.^12,32^ Moreover, our model utilizing human mobility data from Facebook is not limited to intercity or intracity level, or Taiwan. Facebook mobility data are also calculated at finer geographic scales (such as towns) and for other countries, and our model can be easily applied in these settings to understand disease dynamics of COVID-19.

## CONCLUSIONS

In Taiwan, most cases to date were imported or linked to imported cases. To prepare for the potential spread within Taiwan, we utilized Facebook’s aggregated and anonymized movement and colocation data to identify cities with higher risk of infection and regional importation. We showed that both intracity and intercity movement affect outbreak dynamics, with the former having more of an impact on the total numbers of cases and the latter impacting geographic scope. The timing, duration, and level of travel reduction together determine the impact of travel reductions on the number of infections, and multiple combinations of these can result in similar impact. These findings have important implications for guiding future policies for travel restrictions during outbreaks in Taiwan.

## Data Availability

Modeling output data are in the manuscript or can be accessed through the publicly available web-based interface (https://roachchang.shinyapps.io/TW_CoV_Dynamics/). Due to the Data Sharing Agreement with Facebook, readers can not access the original Facebook data used in this study.

## DECLARATIONS

### Ethics approval and consent to participate

This study received ethical approval from Research Ethics Committee of National Tsing Hua University (REC #: 10812HM119). Aggregated, anonymized data were analyzed, and no individuals were enrolled into this study.

### Consent for publication

Not applicable

### Availability of data and materials

Modeling output data are in the manuscript or can be accessed through publicly available web-based interface (https://roachchang.shinyapps.io/TW_CoV_Dynamics/) . Due to the Data Sharing Agreement with Facebook, readers can not access the original Facebook mobility data used in this study.

### Authors’ contributions

HHC, CSL, and COB designed the experiments. HHC and RK created the models. MCC ran the simulations. MCC and HHC performed analysis. HHC, RK, and MCC interpreted the results and wrote the manuscript. YAL and CSL collected data for the models. All authors have read and approved the manuscript.

## Acknowledgements

We thank Brian Karrer, Alex Pompe, and Laura McGorman from Facebook. This study was supported by the Ministry of Science and Technology in Taiwan (MOST 109-2636-B-007-006). MCC, YAL, and HHC were supported by Yushan Scholar Program, CSL was supported by the Ministry of Science and Technology in Taiwan (MOST109-2636-B-007-004), RK was supported by National Institute of General Medical Sciences (U54GM088558), COB was supported by National Institute of General Medical Sciences (R35GM124715-02). The funders had no role in study design, data collection, data analysis, data interpretation, or writing of the report.

**List of Abbreviations**
COVID-19: Coronavirus Disease 2019
SLIR: Susceptible-latent-infectious-recovered

## SUPPLEMENTARY MATERIALS

### SUPPLEMENTARY METHODS

#### Estimating *P_ij_*

We built a travel model to estimate the proportion of time people living in location *i* spend in location *j* (*P_ij_*) by fitting the model to the Facebook movement data. *X_ij_* represents the proportion of people living in location *i* currently in location *j*, and 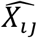 represents the equilibrium state of *X_ij_*, and its value under the fitted model is used as our estimate of *P_ij_*. People living in location *i* travel with probability *F_i_*, and the probability that a traveler from location *i* travels to location *j* is denoted by *T_ij_*. Travelers go back to their home location at probability *λ_i_* per unit of time. *M_ij,t,t+1_* represents the number of people moving from location *i* to location *j* between time *t* and *t*+1.

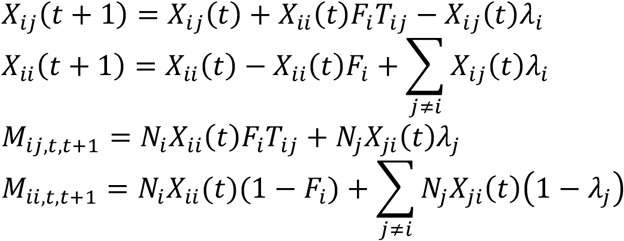

At equilibrium,

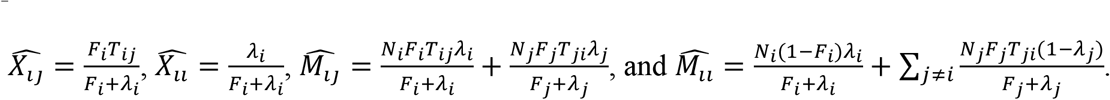

For simplicity, we assumed that the majority of travel is work-related travel and on average travelers spend eight hours in the travel destination (*λ_i_* =1 given the unit of time is 8 hours) and that *T_ij_* is proportional to *M_ij_*, leaving *F_i_* the only parameters to be fitted. We used a gradient descent algorithm to find the local optimum solution for *F_i_*, where the cost function is defined by the sum of the squared difference between normalized *m_ij_* and the normalized value of *M_ij_* from the model. We calculated 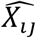 under fitted parameters to obtain estimates of *P_ij_*.

### SUPPLEMENTARY FIGURES

**Figure S1.**
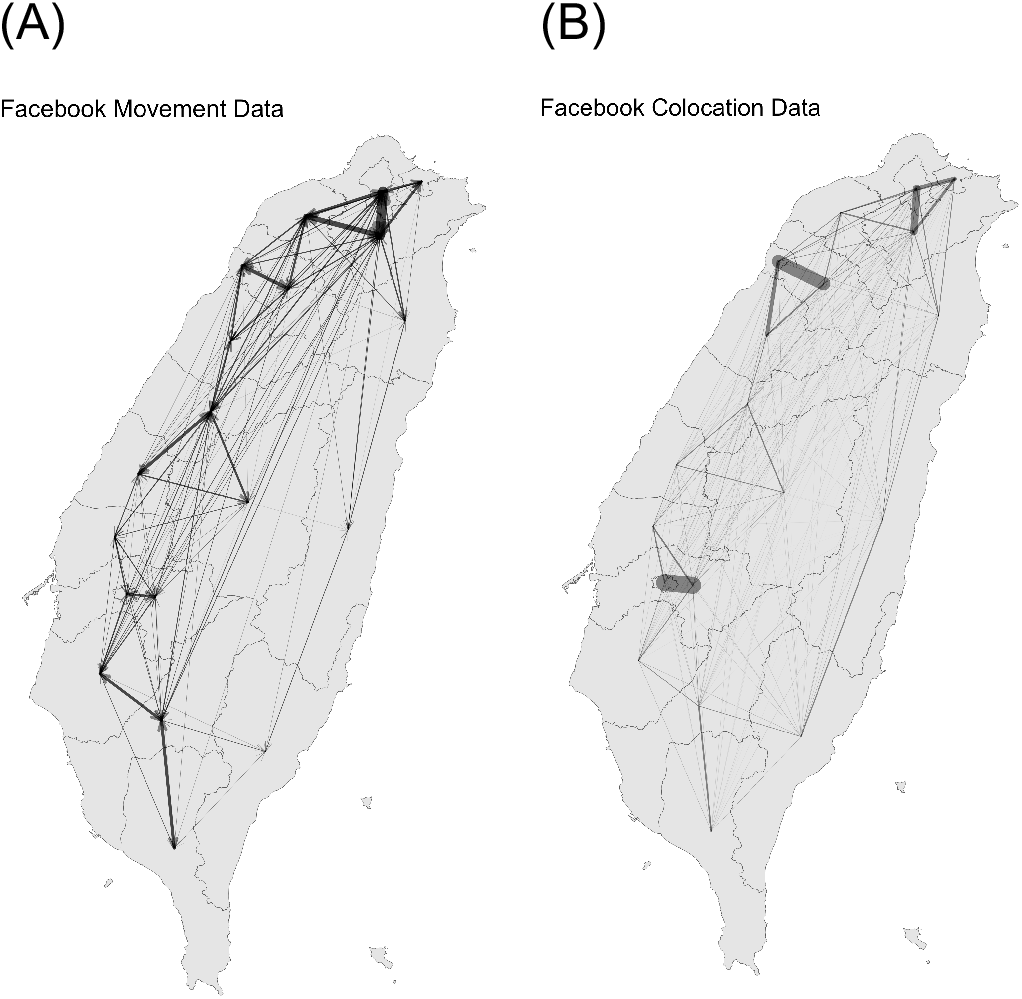
Movement patterns estimated from the Facebook data in Taiwan. (A) Regular movement data. (B) Colocation matrices.

**Figure S2.**
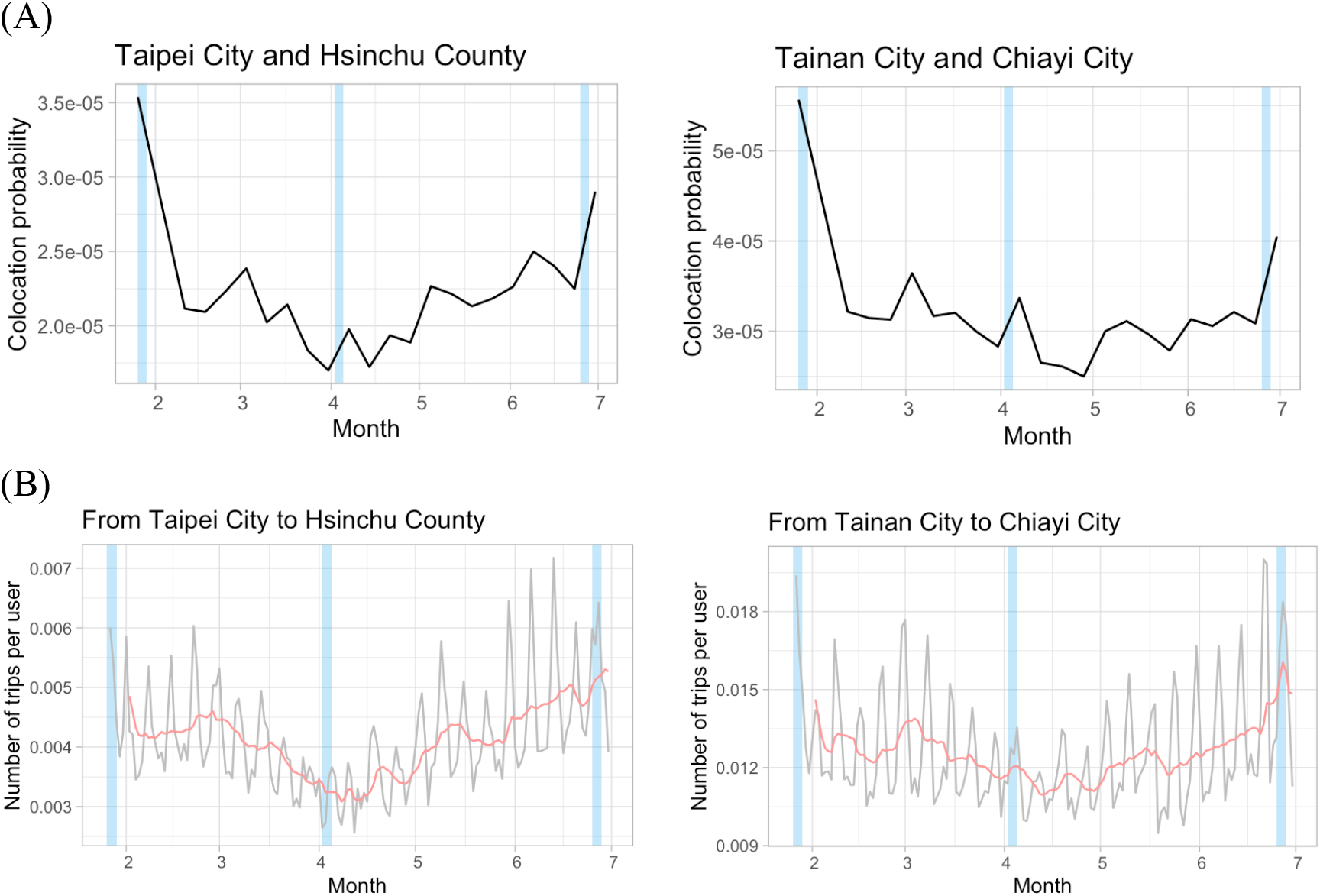
Mobility change over time. Two examples of city pairs where the baseline travel first decreased and then increased between February and June were shown for both (**A**) colocation and (**B**) movement data. The dates of major holidays (Lunar New Year, Ching Ming Festival, and Dragon Boat Festival) are shown in blue.

**Figure S3.**
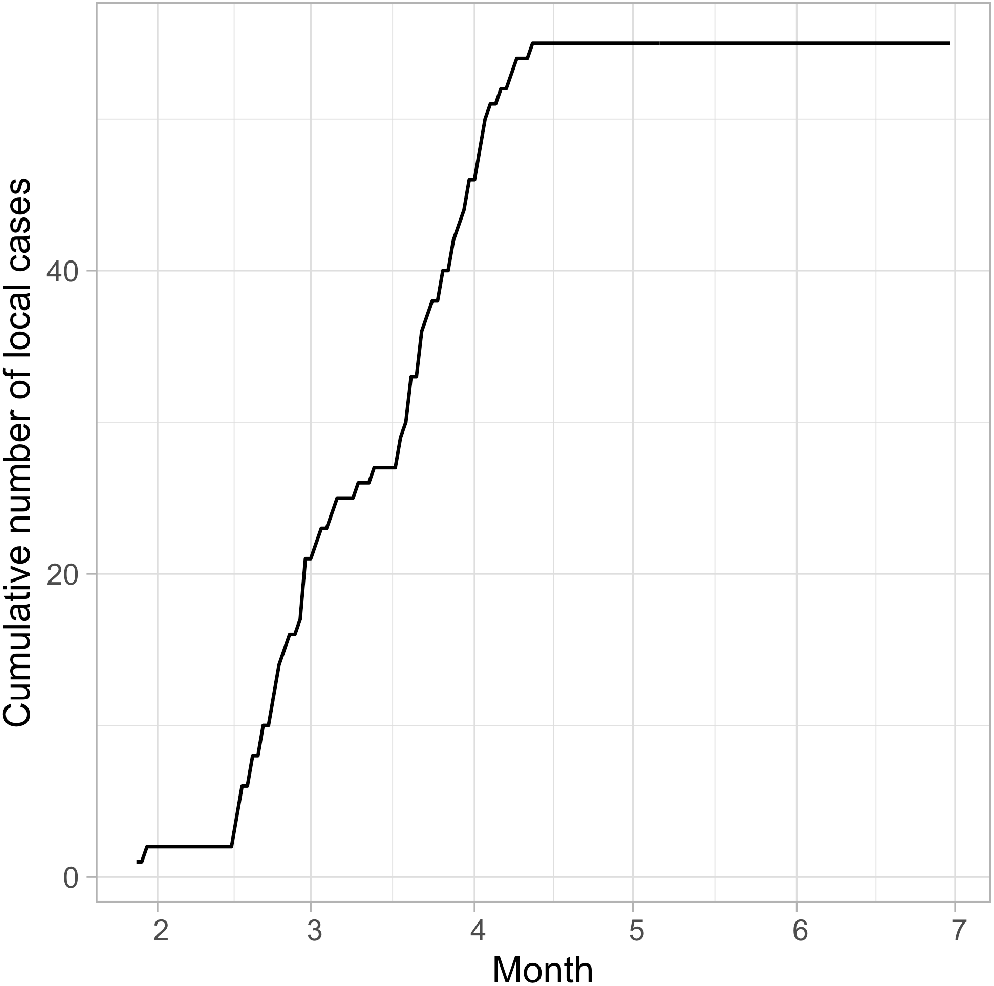
Cumulative number of local cases in Taiwan.

**Figure S4.**
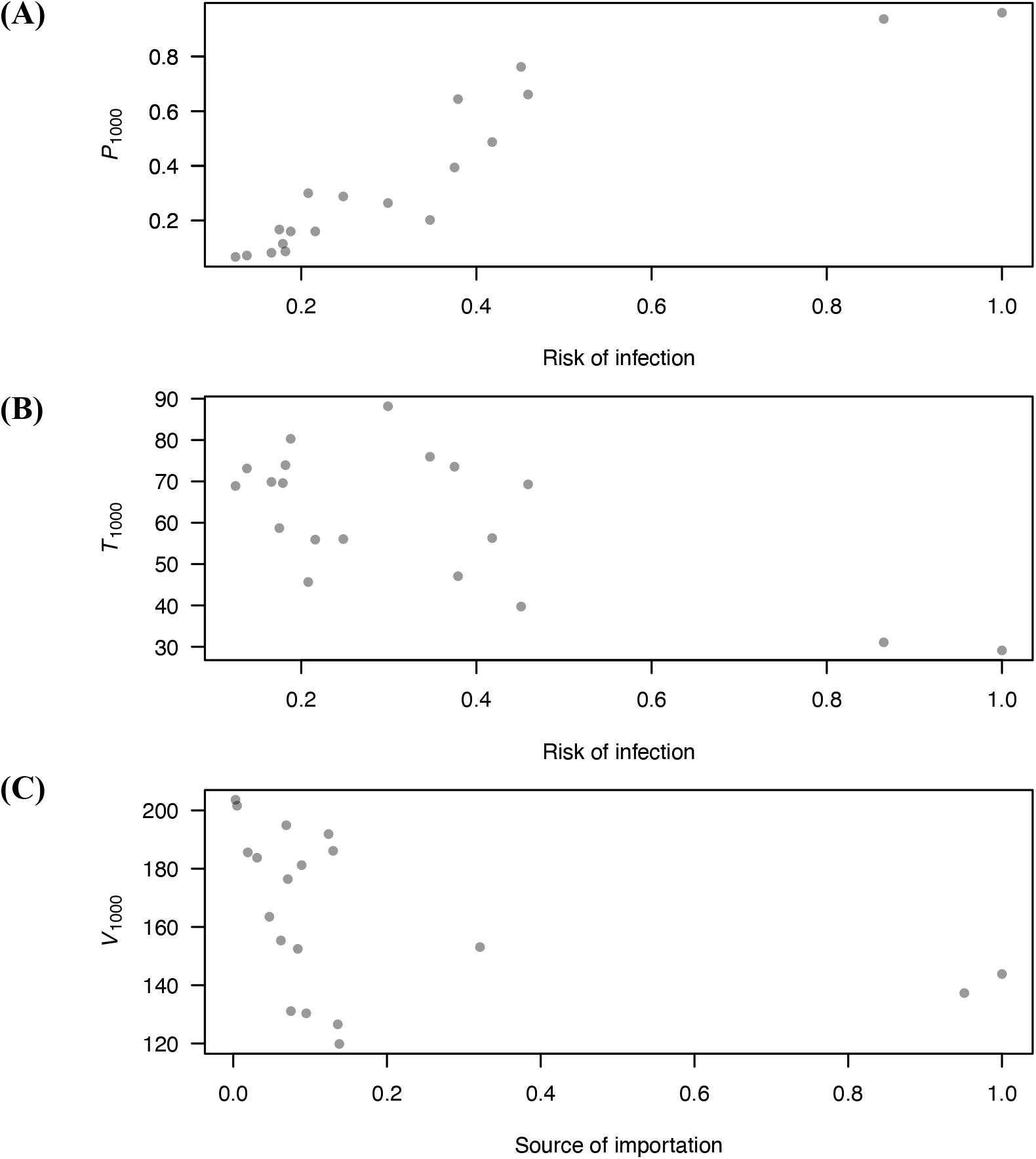
Disease spread was associated with measures of connectivity. In the contact model, (**A**) the probability of having more than 1000 infections (*P_1000_*) increased with risk of infection (Spearman’s correlation test, rho= 0.95, *p*-value=5×10^-10^), and (**B**) the time it took to reach 1000 infections (*T_1000_*) decreased with risk of infection (Spearman’s correlation test, rho= - 0.46, *p*-value=0.05). (**C**) In the residence model, the variation in infection numbers across cities at *T_1000_* (denoted by *V_1000_*) decreased with values of source of importation (Spearman’s correlation test, rho= -0.64, *p*-value=0.004). *R_0_*=2.4.

**Figure S5.**
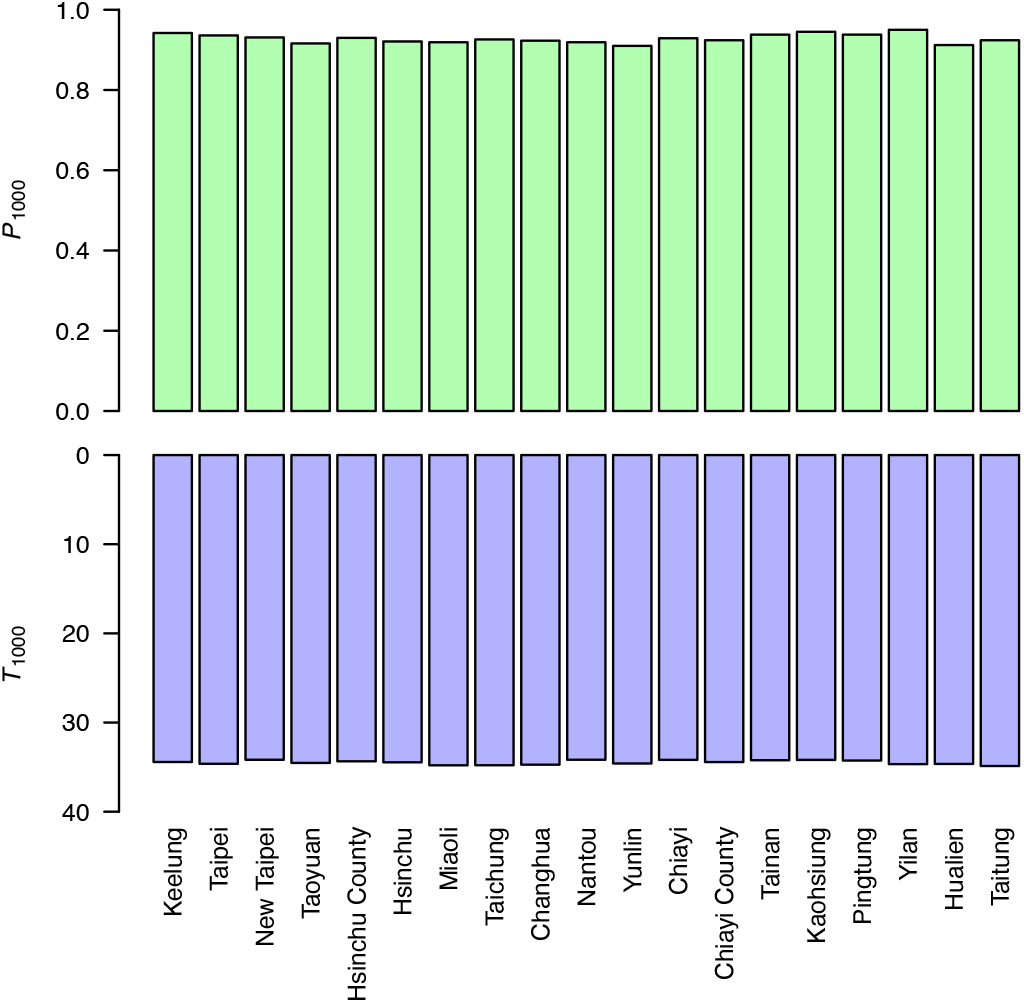
*P_1000_* and *T_1000_* did not vary much with the locations of initial infections. *R_0_*=2.4.

**Figure S6.**
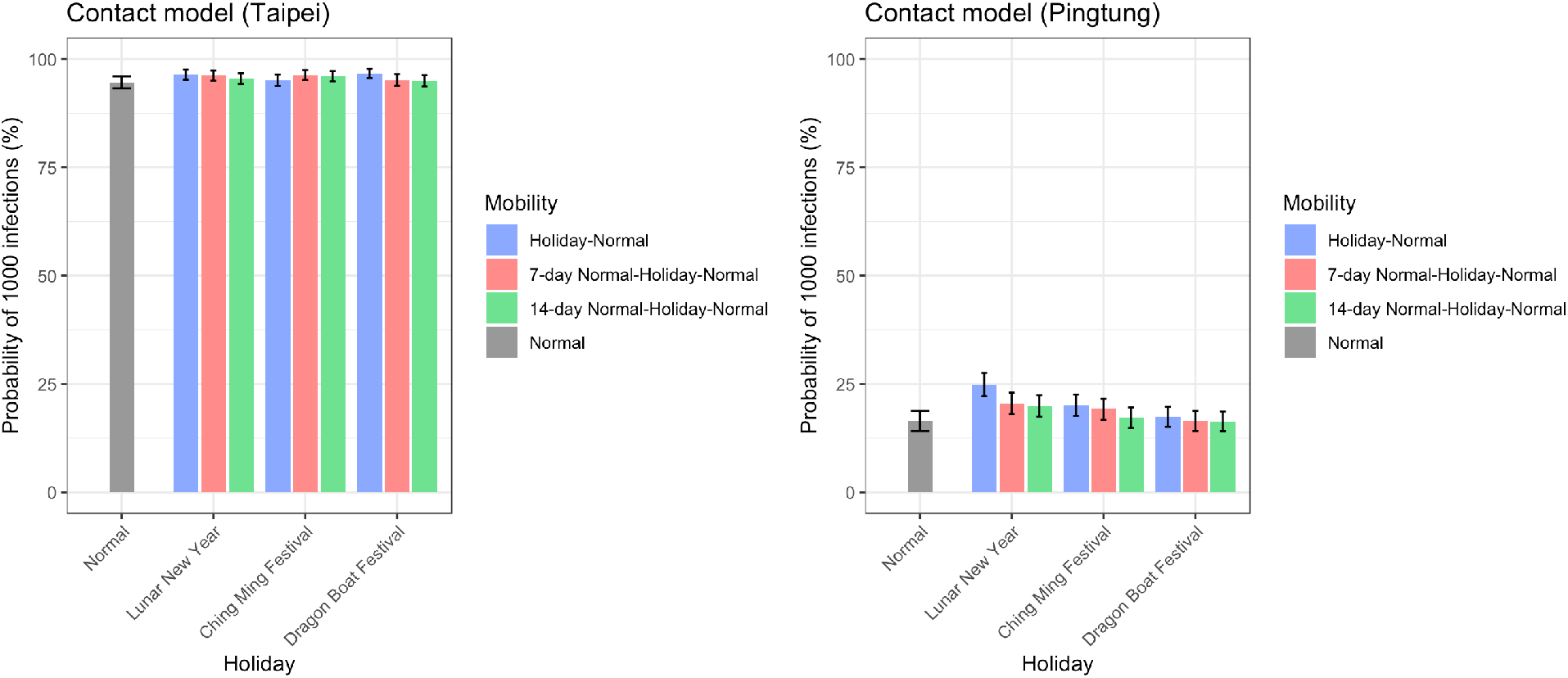
The impact of holiday travel on the probability of outbreak. The probability of outbreak (*P_1000_*) increased with mobility during Lunar New Year (10-day). The impact of Ching Ming Festival (4-day) and Dragon Boat Festival (4-day) is less apparent. Initial infections occurred either in (blue) or before holidays (red: 7-day; green: 14-day). *R_0_*=2.4.

**Figure S7.**
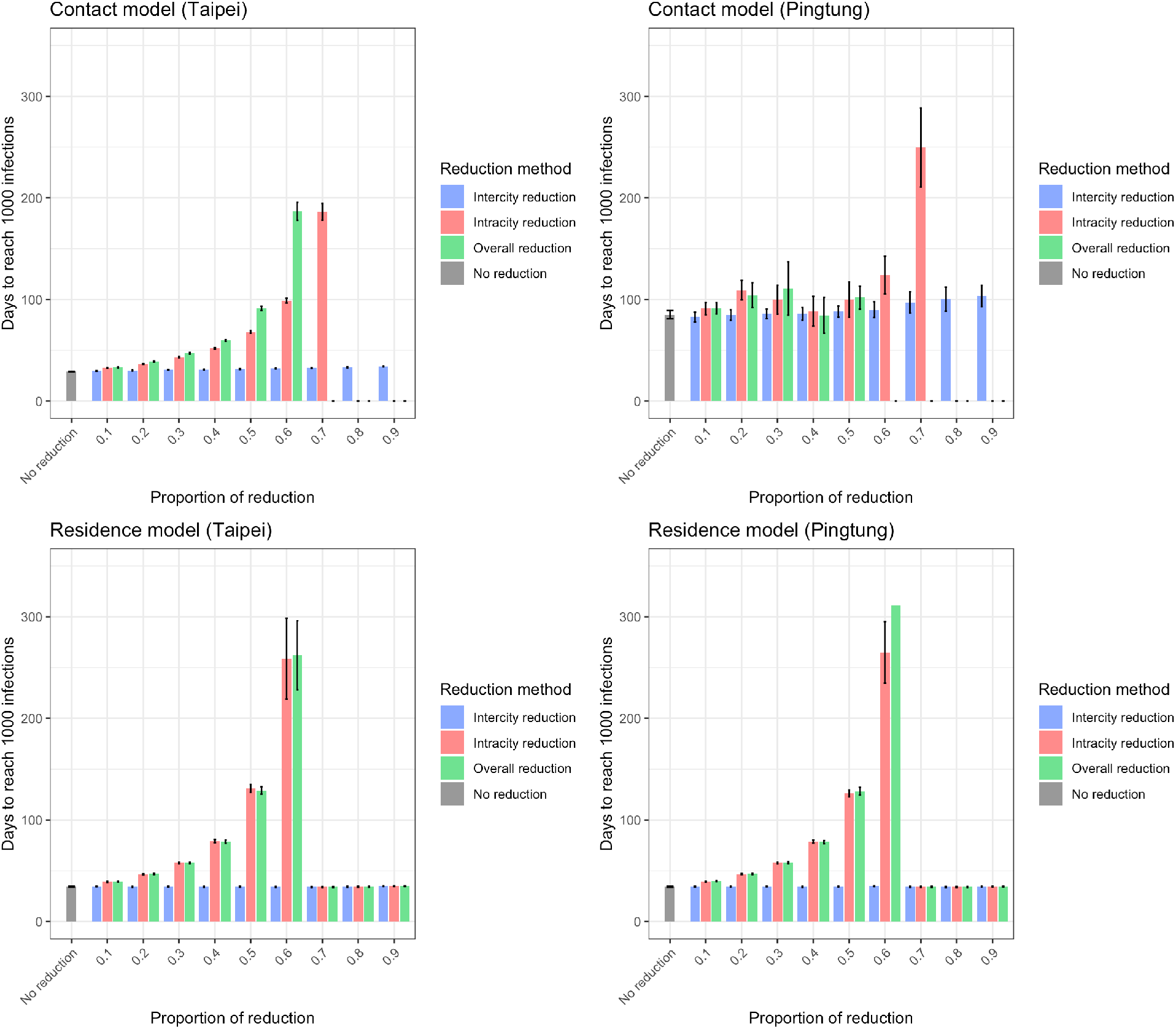
The impact of travel reduction on time to reach 1000 accumulated infections. If initial infections were in a big city, it took less time to reach 1000 infections in the contact model. The difference between big and small cities was not significant in the residence model. Intracity and overall travel reduction delayed the time to reach 1000 infections in both models, while intercity reduction did not. For some conditions, *P_1000,3_* was 0 and no bar was shown. Here travel reduction was applied during the whole time and *R_0_*=2.4.

**Figure S8.**
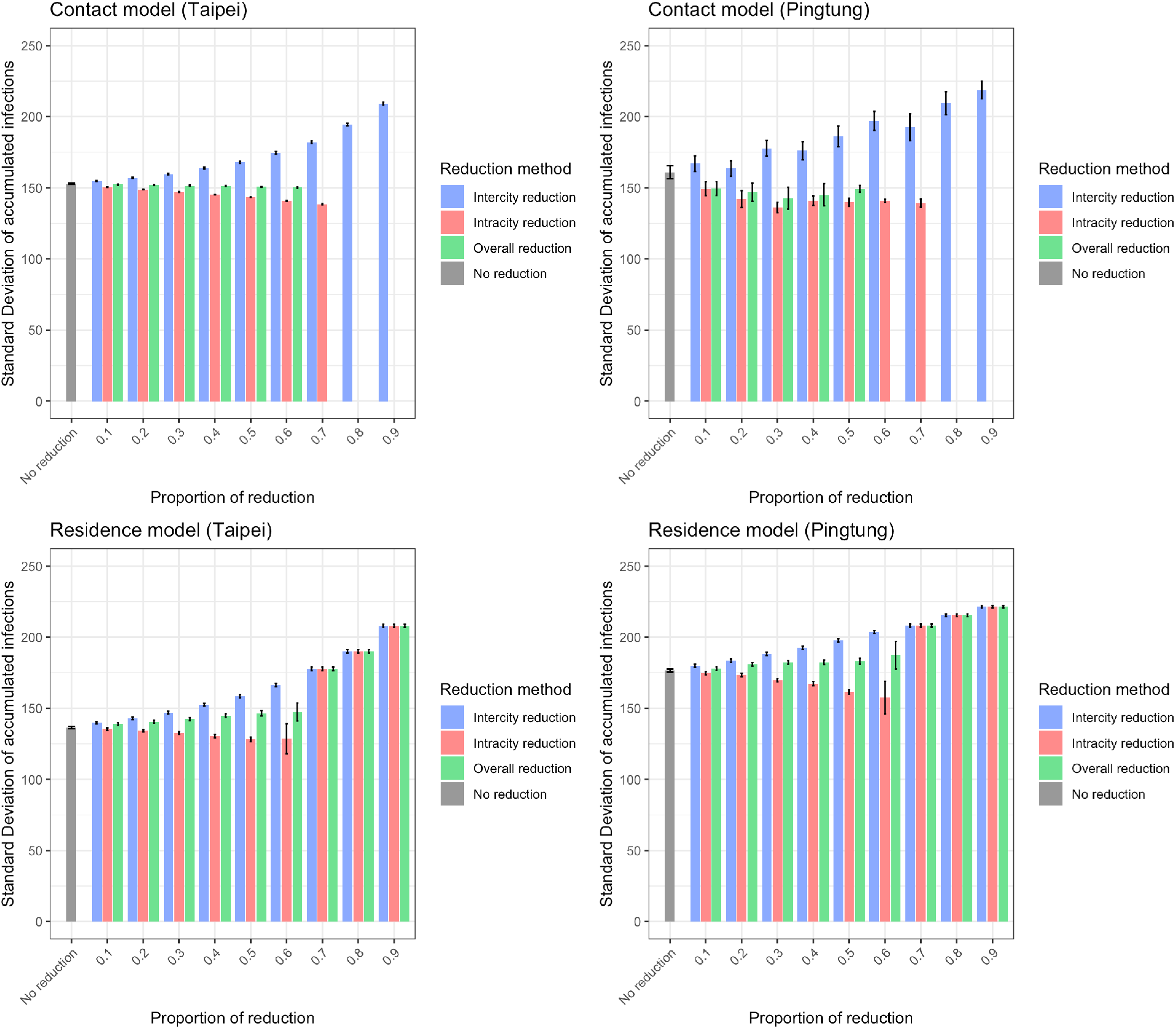
The impact of travel reduction on the geographic distribution of infections. Standard deviation of infection numbers across different cities when there are 1000 infections (*V_1000,3_*) was shown. Intercity travel reduction increased the variation in infection numbers across cities in both models. Here travel reduction was applied during the whole time and *R_0_*=2.4.

**Figure S9.**
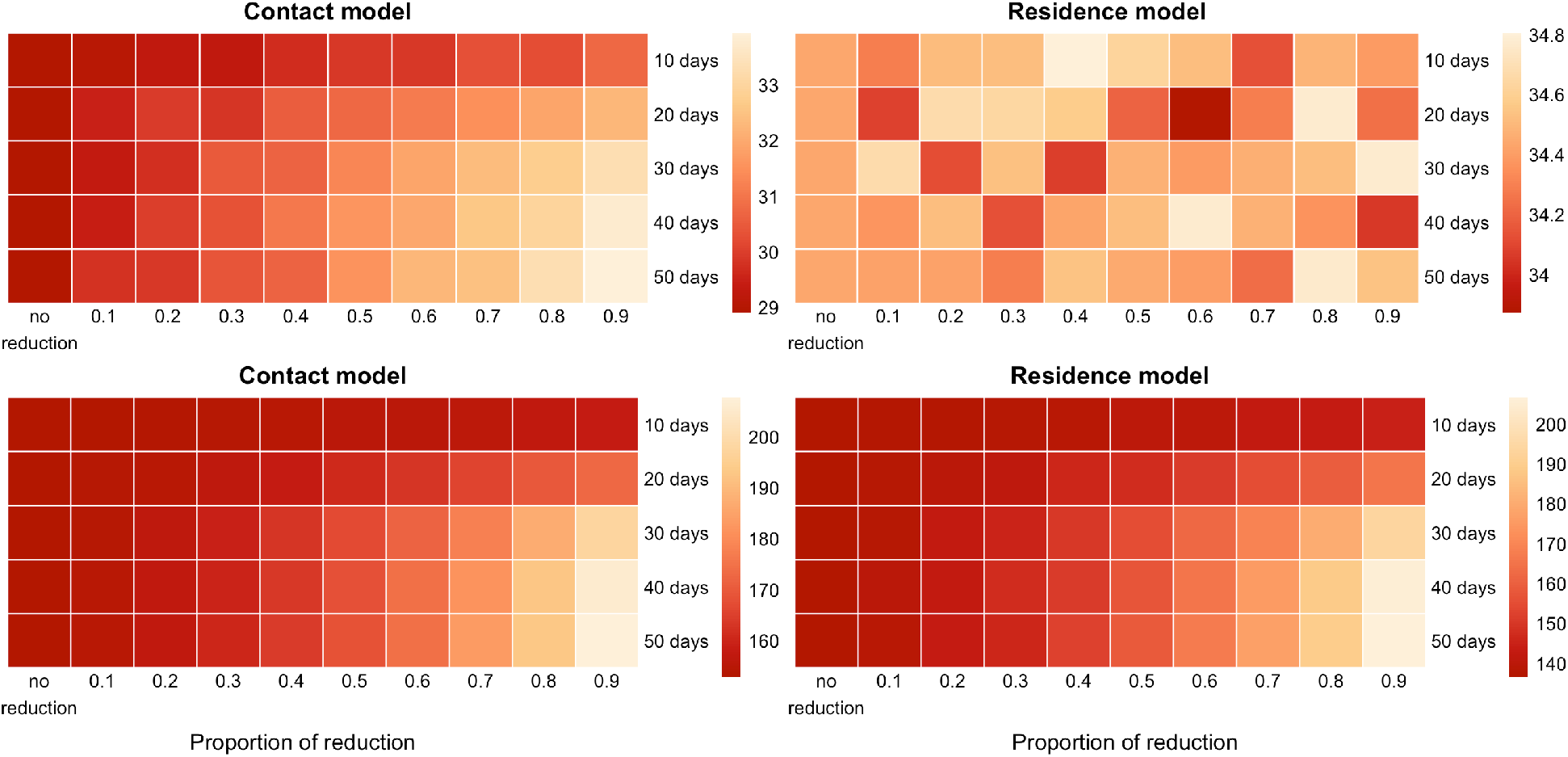
*T_1000,3_* and *V_1000,3_* under different lengths of intercity travel reduction. *T_1000,3_* (upper panel) and *V_1000,3_* (lower panel). Here initial infections were in Taipei city and *R_0_*=2.4.

**Figure S10.**
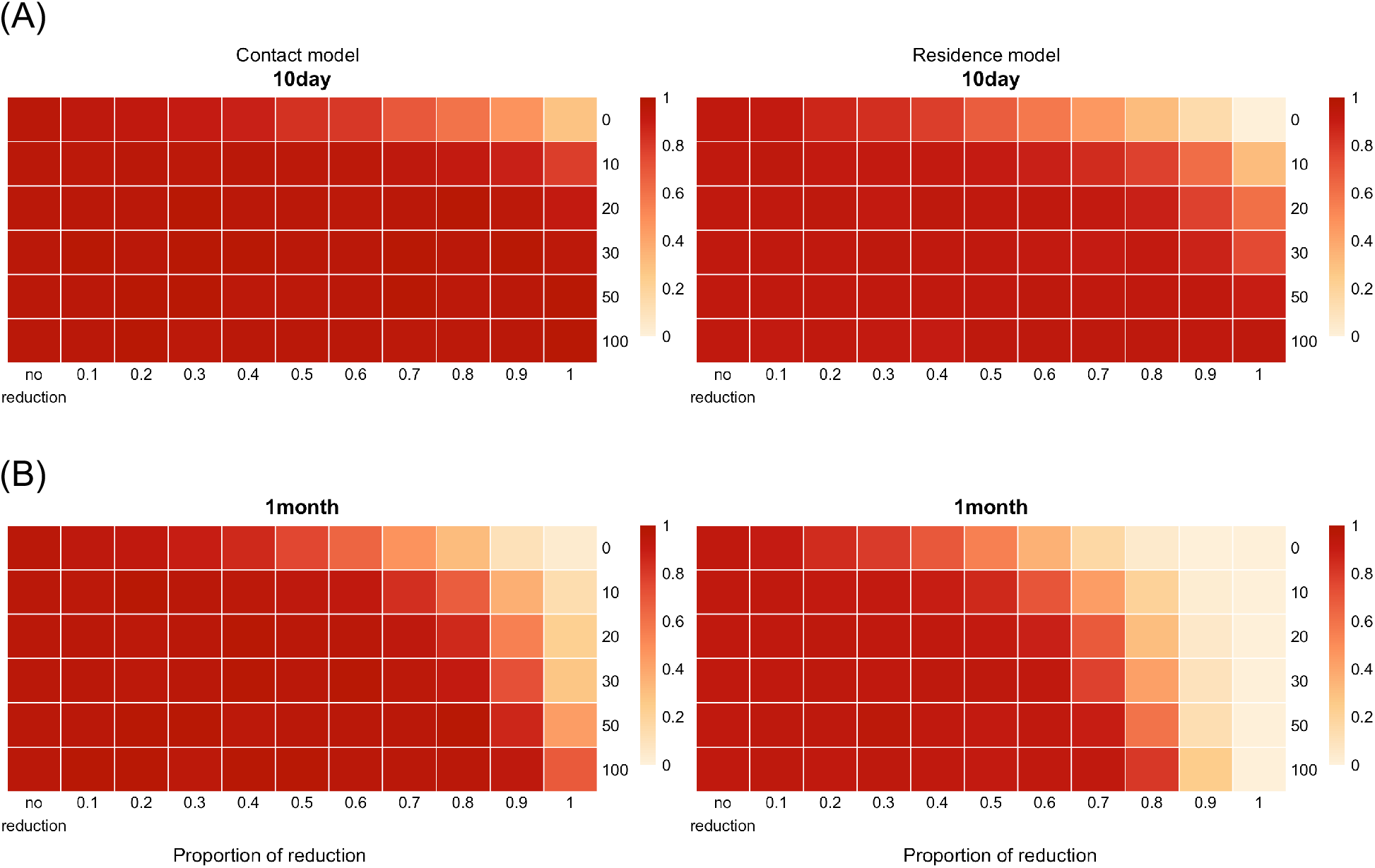
*P_1000,3_* when travel reduction started at different conditions. *P_1000,3_* when travel reduction started from the beginning of the simulations (denoted by 0), or when there were 10, 20, 30, 50, and 100 infections in both contact (left) and residence (right) models. Two different lengths of travel reduction duration were shown: (**A**) 10 days (**B**) 1 month. Only intracity travel reduction was shown here because intercity travel reduction only had minimal impact on *P_1000,3_* and the results from overall reduction and intracity reduction were qualitatively similar. It was best to reduce travel at the beginning if the duration was for 10 days or 1 month. Here initial infections were in Taipei city and *R_0_*=2.4.

### SUPPLEMENTARY TABLES

**Table S1.** Intracity *R_0_*, intercity *R_0_*, risk of infection, and risk of importation.

| City | Intracity<br>$R_0$ | Intercity<br>$R_0$ | Risk of<br>infection | Risk of<br>importation | Source of<br>importation |
| --- | --- | --- | --- | --- | --- |
| Keelung City | 1.016 | **0.348** | **0.451** | 0.107 | 0.095 |
| New Taipei City | **2.247<sup>#</sup>** | **0.368** | **0.865** | **0.125** | **1.000** |
| Taipei City | **2.500** | **0.523** | **1.000** | **0.155** | **0.951** |
| Taoyuan City | 0.985 | 0.162 | 0.379 | 0.073 | **0.321** |
| Hsinchu County | 0.562 | 0.187 | 0.248 | **0.123** | **0.136** |
| Hsinchu City | **1.023** | **0.241** | **0.418** | **0.143** | **0.138** |
| Miaoli County | 0.425 | 0.104 | 0.175 | 0.044 | 0.062 |
| Taichung City | **1.051** | 0.081 | 0.375 | 0.026 | 0.130 |
| Changhua County | 0.475 | 0.074 | 0.182 | 0.029 | 0.089 |
| Yunlin County | 0.348 | 0.071 | 0.138 | 0.019 | 0.031 |
| Chiayi County | 0.258 | 0.121 | 0.125 | 0.073 | 0.084 |
| Chiayi City | 0.836 | **0.212** | 0.347 | **0.141** | 0.075 |
| Nantou County | 0.408 | 0.092 | 0.166 | 0.036 | 0.047 |
| Tainan City | 0.842 | 0.063 | 0.299 | 0.019 | 0.069 |
| Kaohsiung City | **1.323** | 0.066 | **0.459** | 0.022 | 0.124 |
| Pingtung County | 0.482 | 0.087 | 0.188 | 0.036 | 0.071 |
| Taitung County | 0.468 | 0.072 | 0.179 | 0.004 | 0.003 |
| Hualien County | 0.590 | 0.063 | 0.216 | 0.006 | 0.005 |
| Yilan County | 0.524 | 0.106 | 0.208 | 0.018 | 0.019 |

<sup>#</sup>Top five values in each column are bold.

**Table S2.** The probability of having 1000 infections given different numbers of initial infections in different cities (contact model). Colocation matrices in regular days were used. *R_0_*=2.4.

|  | 1 | 2 | 3 | 4 | 5 | 6 | 7 | 8 | 9 | 10 |
| --- | --- | --- | --- | --- | --- | --- | --- | --- | --- | --- |
| **Keelung City** | 0.396 | 0.611 | 0.783 | 0.849 | 0.904 | 0.944 | 0.967 | 0.979 | 0.985 | 0.992 |
| **New Taipei City** | 0.619 | 0.871 | 0.933 | 0.982 | 0.997 | 0.997 | 1.000 | 1.000 | 1.000 | 1.000 |
| **Taipei City** | 0.677 | 0.882 | 0.953 | 0.983 | 0.993 | 0.997 | 1.000 | 1.000 | 1.000 | 1.000 |
| **Taoyuan City** | 0.285 | 0.473 | 0.603 | 0.733 | 0.805 | 0.835 | 0.907 | 0.924 | 0.936 | 0.948 |
| **Hsinchu County** | 0.102 | 0.207 | 0.264 | 0.342 | 0.405 | 0.487 | 0.575 | 0.603 | 0.649 | 0.676 |
| **Hsinchu City** | 0.203 | 0.354 | 0.503 | 0.636 | 0.713 | 0.748 | 0.789 | 0.827 | 0.865 | 0.908 |
| **Miaoli County** | 0.058 | 0.103 | 0.160 | 0.185 | 0.233 | 0.287 | 0.318 | 0.362 | 0.388 | 0.397 |
| **Taichung City** | 0.162 | 0.302 | 0.394 | 0.492 | 0.565 | 0.620 | 0.708 | 0.735 | 0.800 | 0.824 |
| **Changhua County** | 0.032 | 0.066 | 0.090 | 0.115 | 0.152 | 0.158 | 0.201 | 0.225 | 0.256 | 0.285 |
| **Yunlin County** | 0.020 | 0.038 | 0.064 | 0.105 | 0.116 | 0.141 | 0.160 | 0.183 | 0.197 | 0.220 |
| **Chiayi County** | 0.031 | 0.057 | 0.089 | 0.097 | 0.128 | 0.145 | 0.158 | 0.179 | 0.207 | 0.230 |
| **Chiayi City** | 0.084 | 0.144 | 0.217 | 0.272 | 0.303 | 0.402 | 0.434 | 0.479 | 0.533 | 0.554 |
| **Nantou County** | 0.026 | 0.057 | 0.102 | 0.115 | 0.139 | 0.183 | 0.193 | 0.214 | 0.245 | 0.279 |
| **Tainan City** | 0.104 | 0.170 | 0.244 | 0.353 | 0.389 | 0.467 | 0.482 | 0.540 | 0.559 | 0.619 |
| **Kaohsiung City** | 0.310 | 0.557 | 0.678 | 0.781 | 0.858 | 0.906 | 0.927 | 0.961 | 0.971 | 0.980 |
| **Pingtung County** | 0.042 | 0.103 | 0.166 | 0.240 | 0.280 | 0.299 | 0.317 | 0.371 | 0.419 | 0.461 |
| **Taitung County** | 0.042 | 0.073 | 0.109 | 0.141 | 0.173 | 0.204 | 0.216 | 0.269 | 0.283 | 0.298 |
| **Hualien County** | 0.070 | 0.142 | 0.180 | 0.251 | 0.281 | 0.324 | 0.372 | 0.392 | 0.470 | 0.429 |
| **Yilan County** | 0.096 | 0.188 | 0.283 | 0.326 | 0.406 | 0.453 | 0.491 | 0.558 | 0.622 | 0.651 |

**Table S3.** The probability of having 1000 infections given different numbers of initial infections in different cities (residence model). Movement data on weekdays were used. *R_0_*=2.4.

|  | 1 | 2 | 3 | 4 | 5 | 6 | 7 | 8 | 9 | 10 |
| --- | --- | --- | --- | --- | --- | --- | --- | --- | --- | --- |
| **Keelung City** | 0.571 | 0.820 | 0.942 | 0.962 | 0.988 | 0.997 | 0.994 | 0.998 | 0.999 | 1.000 |
| **New Taipei City** | 0.593 | 0.835 | 0.923 | 0.968 | 0.987 | 0.992 | 1.000 | 1.000 | 1.000 | 1.000 |
| **Taipei City** | 0.646 | 0.842 | 0.926 | 0.973 | 0.989 | 0.996 | 0.998 | 0.999 | 1.000 | 0.999 |
| **Taoyuan City** | 0.614 | 0.826 | 0.922 | 0.965 | 0.988 | 0.995 | 0.999 | 0.999 | 1.000 | 1.000 |
| **Hsinchu County** | 0.587 | 0.824 | 0.933 | 0.956 | 0.984 | 0.996 | 0.997 | 1.000 | 1.000 | 1.000 |
| **Hsinchu City** | 0.615 | 0.821 | 0.910 | 0.975 | 0.993 | 0.997 | 1.000 | 1.000 | 1.000 | 1.000 |
| **Miaoli County** | 0.612 | 0.824 | 0.932 | 0.967 | 0.989 | 0.994 | 0.996 | 1.000 | 0.999 | 1.000 |
| **Taichung City** | 0.594 | 0.807 | 0.923 | 0.975 | 0.987 | 0.993 | 0.996 | 0.998 | 1.000 | 1.000 |
| **Changhua County** | 0.590 | 0.841 | 0.925 | 0.970 | 0.993 | 0.996 | 0.999 | 0.999 | 1.000 | 1.000 |
| **Yunlin County** | 0.583 | 0.833 | 0.938 | 0.964 | 0.987 | 0.998 | 0.996 | 1.000 | 1.000 | 1.000 |
| **Chiayi County** | 0.589 | 0.804 | 0.924 | 0.964 | 0.988 | 0.996 | 0.998 | 1.000 | 1.000 | 0.999 |
| **Chiayi City** | 0.568 | 0.848 | 0.937 | 0.967 | 0.993 | 0.995 | 0.997 | 1.000 | 1.000 | 1.000 |
| **Nantou County** | 0.587 | 0.810 | 0.927 | 0.975 | 0.982 | 0.993 | 1.000 | 0.999 | 1.000 | 1.000 |
| **Tainan City** | 0.563 | 0.830 | 0.934 | 0.969 | 0.987 | 0.993 | 0.998 | 0.999 | 0.999 | 1.000 |
| **Kaohsiung City** | 0.561 | 0.834 | 0.913 | 0.975 | 0.987 | 0.996 | 0.998 | 0.999 | 1.000 | 1.000 |
| **Pingtung County** | 0.565 | 0.844 | 0.939 | 0.969 | 0.989 | 0.995 | 1.000 | 1.000 | 1.000 | 1.000 |
| **Taitung County** | 0.616 | 0.843 | 0.922 | 0.964 | 0.982 | 0.993 | 0.998 | 1.000 | 1.000 | 1.000 |
| **Hualien County** | 0.574 | 0.836 | 0.922 | 0.955 | 0.992 | 0.994 | 0.998 | 0.999 | 1.000 | 1.000 |
| **Yilan County** | 0.586 | 0.823 | 0.928 | 0.979 | 0.985 | 0.997 | 0.997 | 0.999 | 1.000 | 1.000 |

## REFERENCES

1. Dong E, Du H, Gardner L. An interactive web-based dashboard to track COVID-19 in real time. Lancet Infect Dis. 2020;20(5):533–4.

2. Maxmen A. More than 80 clinical trials launch to test coronavirus treatments. Nature. 2020;578(7795):347.

3. STAT. Covid-19 Drugs & Vaccine Tracker [Internet]. STAT News. 2020. Available from: https://www.statnews.com/feature/coronavirus/drugs-vaccines-tracker/

4. Ferguson NM, Laydon D, Nedjati-gilani G, Imai N, Ainslie K, Baguelin M, et al. Impact of non-pharmaceutical interventions (NPIs) to reduce COVID- 19 mortality and healthcare demand. 2020;(March).

5. Kissler SM, Tedijanto C, Goldstein E, Grad YH, Lipsitch M. Projecting the transmission dynamics of SARS-CoV-2 through the postpandemic period. Science (80-) [Internet]. 2020 May 22;368(6493):860 LP – 868. Available from: http://science.sciencemag.org/content/368/6493/860.abstract

6. Chinazzi M, Davis JT, Ajelli M, Gioannini C, Litvinova M, Merler S, et al. The effect of travel restrictions on the spread of the 2019 novel coronavirus (COVID-19) outbreak. Science (80-). 2020;

7. Bhatia S, Imai N, Cuomo-dannenburg G, Baguelin M, Boonyasiri A, Cori A, et al. Report 6: Relative sensitivity of international surveillance. 2020;(Figure 1):1-6.

8. Niehus R, De Salazar PM, Taylor AR, Lipsitch M. Using observational data to quantify bias of traveller-derived COVID-19 prevalence estimates in Wuhan, China. Lancet Infect Dis. 2020;

9. Taiwan CDC. COVID-19.

10. Bureau of Consular Affairs, Ministry of Foreign Affairs R of C (Taiwan). News & Events [Internet]. Available from: https://www.boca.gov.tw/lp-220-2.html

11. World Health Organization. Responding to community spread of COVID-19. 2020;(March): 1-6.

12. Buckee CO, Balsari S, Chan J, Crosas M, Dominici F, Gasser U, et al. Aggregated mobility data could help fight COVID-19. Science (80- ) [Internet]. 2020 Mar 23;eabb8021. Available from: http://science.sciencemag.org/content/early/2020/03/20/science.abb8021.abstract

13. Li R, Pei S, Chen B, Song Y, Zhang T, Yang W, et al. Substantial undocumented infection facilitates the rapid dissemination of novel coronavirus (SARS-CoV2). Science (80- ) [Internet]. 2020 Mar 16;eabb3221. Available from: http://science.sciencemag.org/content/early/2020/03/13/science.abb3221.abstract

14. Kraemer MUG, Yang C-H, Gutierrez B, Wu C-H, Klein B, Pigott DM, et al. The effect of human mobility and control measures on the COVID-19 epidemic in China. Science (80- ) [Internet]. 2020 Mar 25;eabb4218. Available from: http://science.sciencemag.org/content/early/2020/03/25/science.abb4218.abstract

15. Gatto M, Bertuzzo E, Mari L, Miccoli S, Carraro L, Casagrandi R, et al. Spread and dynamics of the COVID-19 epidemic in Italy: Effects of emergency containment measures. Proc Natl Acad Sci [Internet]. 2020 May 12;117(19): 10484 LP - 10491. Available from: http://www.pnas.org/content/117/19/10484.abstract

16. Kissler S, Kishore N, Prabhu M, Goffman D, Beilin Y, Landau R, et al. Reductions in commuting mobility predict geographic differences in SARS-CoV-2 prevalence in New York City. 2020;

17. Maas P, Nayak C, Dow A, Gros A, Mason W, Filiz IO, et al. Facebook Disaster Maps: Methodology. Faceb Res [Internet]. 2017;1-10. Available from: https://research.fb.com/facebook-disaster-maps-methodology/

18. Davies NG, Kucharski AJ, Eggo RM, Gimma A, Edmunds WJ, Jombart T, et al. Effects of non-pharmaceutical interventions on COVID-19 cases, deaths, and demand for hospital services in the UK: a modelling study. Lancet Public Heal. 2020;

19. Ferguson N, Laydon D, Nedjati Gilani G, Imai N, Ainslie K, Baguelin M, et al. Report 9: Impact of non-pharmaceutical interventions (NPIs) to reduce COVID19 mortality and healthcare demand. 2020;

20. Ruktanonchai NW, DeLeenheer P, Tatem AJ, Alegana VA, Caughlin TT, zu Erbach-Schoenberg E, et al. Identifying Malaria Transmission Foci for Elimination Using Human Mobility Data. PLOS Comput Biol [Internet]. 2016 Apr 4;12(4):e1004846. Available from: https://doi.org/10.1371/journal.pcbi.1004846

21. Acevedo MA, Prosper O, Lopiano K, Ruktanonchai N, Caughlin TT, Martcheva M, et al. Spatial heterogeneity, host movement and mosquito-borne disease transmission. PLoS One [Internet]. 2015 Jun 1;10(6):e0127552-e0127552. Available from: https://pubmed.ncbi.nlm.nih.gov/26030769

22. Cosner C, Beier JC, Cantrell RS, Impoinvil D, Kapitanski L, Potts MD, et al. The effects of human movement on the persistence of vector-borne diseases. J Theor Biol [Internet]. 2009/03/03. 2009 Jun 21;258(4):550–60. Available from: https://pubmed.ncbi.nlm.nih.gov/19265711

23. Rodríguez DJ, Torres-Sorando L. Models of infectious diseases in spatially heterogeneous environments. Bull Math Biol [Internet]. 2001;63(3):547–71. Available from: https://doi.org/10.1006/bulm.2001.0231

24. McCallum H, Barlow N, Hone J. How should pathogen transmission be modelled? Trends Ecol Evol. 2001;16(6):295–300.

25. Wu JT, Leung K, Leung GM. Nowcasting and forecasting the potential domestic and international spread of the 2019-nCoV outbreak originating in Wuhan, China: a modelling study. Lancet. 2020;395(10225):689–97.

26. Lai S, Ruktanonchai NW, Zhou L, Prosper O, Luo W, Floyd JR, et al. Effect of non-pharmaceutical interventions to contain COVID-19 in China. Nature [Internet]. 2020; Available from: https://doi.org/10.1038/s41586-020-2293-x

27. Prem K, Liu Y, Russell TW, Kucharski AJ, Eggo RM, Davies N, et al. The effect of control strategies to reduce social mixing on outcomes of the COVID-19 epidemic in Wuhan, China: a modelling study. Lancet Public Heal. 2020;

28. Anderson RM, Heesterbeek H, Klinkenberg D, Hollingsworth TD. How will country-based mitigation measures influence the course of the COVID-19 epidemic? Lancet. 2020;395(10228):931–4.

29. Cowling BJ, Chan K-H, Fang VJ, Cheng CKY, Fung ROP, Wai W, et al. Facemasks and hand hygiene to prevent influenza transmission in households: a cluster randomized trial. Ann Intern Med. 2009;151(7):437–46.

30. Tracht SM, Del Valle SY, Hyman JM. Mathematical Modeling of the Effectiveness of Facemasks in Reducing the Spread of Novel Influenza A (H1N1). PLoS One [Internet]. 2010 Feb 10;5(2):e9018. Available from: https://doi.org/10.1371/journal.pone.0009018

31. Jarvis CI, Kv Z, Gimma A. Impact of physical distance measures on transmission in the UK. C Repos. 2020;

32. Emanuele Pepe, Paolo Bajardi, Laetitia Gauvin, Filippo Privitera, Ciro Cattuto MT. COVID-19 outbreak response: first assessment of mobility changes in Italy following lockdown [Internet]. 2020. Available from: https://covid19mm.github.io/in-progress/2020/03/13/first-report-assessment.html

